# Identification of SARS-CoV-2 RNA in Healthcare Heating, Ventilation, and Air Conditioning Units

**DOI:** 10.1101/2020.06.26.20141085

**Authors:** Patrick F. Horve, Leslie Dietz, Mark Fretz, David A. Constant, Andrew Wilkes, John M. Townes, Robert G. Martindale, William B. Messer, Kevin G. Van Den Wymelenberg

**Author notes:** **Corresponding Author(s):** Kevin Van Den Wymelenberg, Biology and the Built Environment Center, University of Oregon, 5231 University of Oregon, Eugene, OR, 97403-5231 Patrick Finn Horve, (541)346-5647, Biology and the Built Environment Center, University of Oregon, 5231 University of Oregon, Eugene, OR, 97403-5231. **Author Contributions:** PFH, LGD, MF, and KVDW conceived of the project scope and MF, JMT, RGM, WBM, and KVDW oversaw project and manuscript development. AW provided significant technical knowledge and access to hospital HVAC systems and air handling units. PFH, LD, and DAC performed sample collection, and initial processing. PFH and LD performed sample transport, nucleic acid isolation, and nucleic acid analysis. PFH performed analysis of raw abundance data and contributed to figure production. MF and AW developed Figure 1. PFH and LD wrote the initial manuscript and KVDW, MF, JMT, WBM, DAC, RGM, and AW provided significant edits to the manuscript.

## Abstract

Available information on Severe Acute Respiratory Syndrome Coronavirus 2 (SARS-CoV-2) transmission by small particle aerosols continues to evolve rapidly. To assess the potential role of heating, ventilation, and air conditioning (HVAC) systems in airborne viral transmission, this study sought to determine the viral presence, if any, on air handling units in a healthcare setting where Coronavirus Disease 2019 (COVID-19) patients were being treated. The presence of SARS-CoV-2 RNA was detected in approximately 25% of samples taken from nine different locations in multiple air handlers. While samples were not evaluated for viral infectivity, the presence of viral RNA in air handlers raises the possibility that viral particles can enter and travel within the air handling system of a hospital, from room return air through high efficiency MERV-15 filters and into supply air ducts. Although no known transmission events were determined to be associated with these specimens, the findings suggest the potential for HVAC systems to facilitate transmission by environmental contamination via shared air volumes with locations remote from areas where infected persons reside. More work is needed to further evaluate the risk of SARS-CoV-2 transmission via HVAC systems and to verify effectiveness of building operations mitigation strategies for the protection of building occupants. These results are important within and outside of healthcare settings and may present a matter of some urgency for building operators of facilities that are not equipped with high-efficiency filtration.

## Introduction

Since its emergence in late 2019, SARS-CoV-2 has spread across the globe and led to the deaths of over 450,000 individuals^1^. The main mechanism of transmission has been identified as respiratory droplet transmission by symptomatic or asymptomatic persons^2–6^. Despite the identification of droplet spread as the most common mechanism of transmission^6^, recent studies suggest that air movement patterns indoors induced through heating, ventilation, and air conditioning (HVAC) systems may contribute to transmission events^7,8^.

Aerosolized SARS-CoV-2 RNA has been previously detected in the air of hospital rooms with symptomatic COVID-19 patients^9–11^, suggesting the possibility that SARS-CoV-2 viral RNA (and potentially virus) have the capacity to enter into building HVAC systems in evacuated room air after a shedding or aerosolization event from infected individuals. Although hospitals contain higher levels of mechanical filtration and room air exchange than almost all other buildings, which are important strategies to help prevent the transmission of disease, a growing body of evidence suggests that these precautions may not be adequate to completely eliminate SARS-CoV-2^12–14^ in filtered air. Studies have shown the persistence of SARS-CoV-2 in air to be hours and on surfaces, days^15^. Efforts to limit the transmission and continued spread of SARS-CoV-2 have mainly focused on social (spatial) distancing, increased cleaning regimens, mandated face coverings, and increased surveillance^15,16^. However, as more indoor spaces begin to reopen and increase in occupant density, more individuals will occupy shared air spaces serviced by HVAC units for extended periods of time. Without an understanding of the risk posed from recirculated air in these indoor environments, there are potential gaps in the prevention and mitigation plans aimed to reduce SARS-CoV-2 transmission and thereby the number of COVID-19 cases.

In the past, ventilation has played a key role in the transmission of infectious disease^8,17–19^. With the demonstration that ventilation systems may contribute to occupant transmission events, HVAC and ventilation guidelines and recommendations have been modified^20,21^. As the knowledge regarding ventilation during the SARS-CoV-2 pandemic continues to expand, it is likely that building operations best practices will continue to be updated in response to that body of knowledge. Here, we present data demonstrating the presence of SARS-CoV-2 RNA at several locations along mechanical ventilation air return and supply pathways, including multiple locations in air handling units (AHUs).

## Materials and Methods

### Sample Collection

Samples were collected from Oregon Health and Science University (OHSU) hospital in Portland, Oregon, USA on four sampling days in May and June 2020. Environmental sampling does not require Institutional Review Board (IRB) approval; however, the project was reviewed by the OHSU IRB and an IRB Exemption was granted for this work.

During May and June 2020, samples were collected from three separate AHUs (Figure 1a). Within each AHU, three areas along the path of airflow were sampled, including the pre-filters, final filters, and supply air dampers (Figure 1b). The pre-filters are rated at MERV10 and final filters are rated at MERV15, both in excess of minimum code requirements^22^. Based upon engineering calculations and equipment documentation available, the HVAC system is capable of cycling air from the ward, to the AHU, and back to the ward in a time between 90 seconds and five minutes, depending on travel distance to room location. Samples were collected using Puritan PurFlock Ultra swabs (catalog #25-3606-U) and swabs were taken in triplicate at each AHU location from the left, middle, and right side of each area along the path of airflow. Swabs were pre-moistened using viral transport media (RMBIO). Swabbing occurred for 20 seconds on an area approximately 20 × 30 cm at each location and swabs were immediately placed into 15 mL conical tubes (Cole-Parmer, catalog #UX-06336-89) containing 1.5 mL viral transport media and stored on ice for transport to a BSL-2 laboratory with enhanced precautions (BSL2+) lab for processing, which typically occurred within two hours after collection. Samples were collected by the same researcher each sampling time, and the researcher did not demonstrate any symptoms of COVID-19 and tested negative by qRT-PCR.

**Figure 1:**
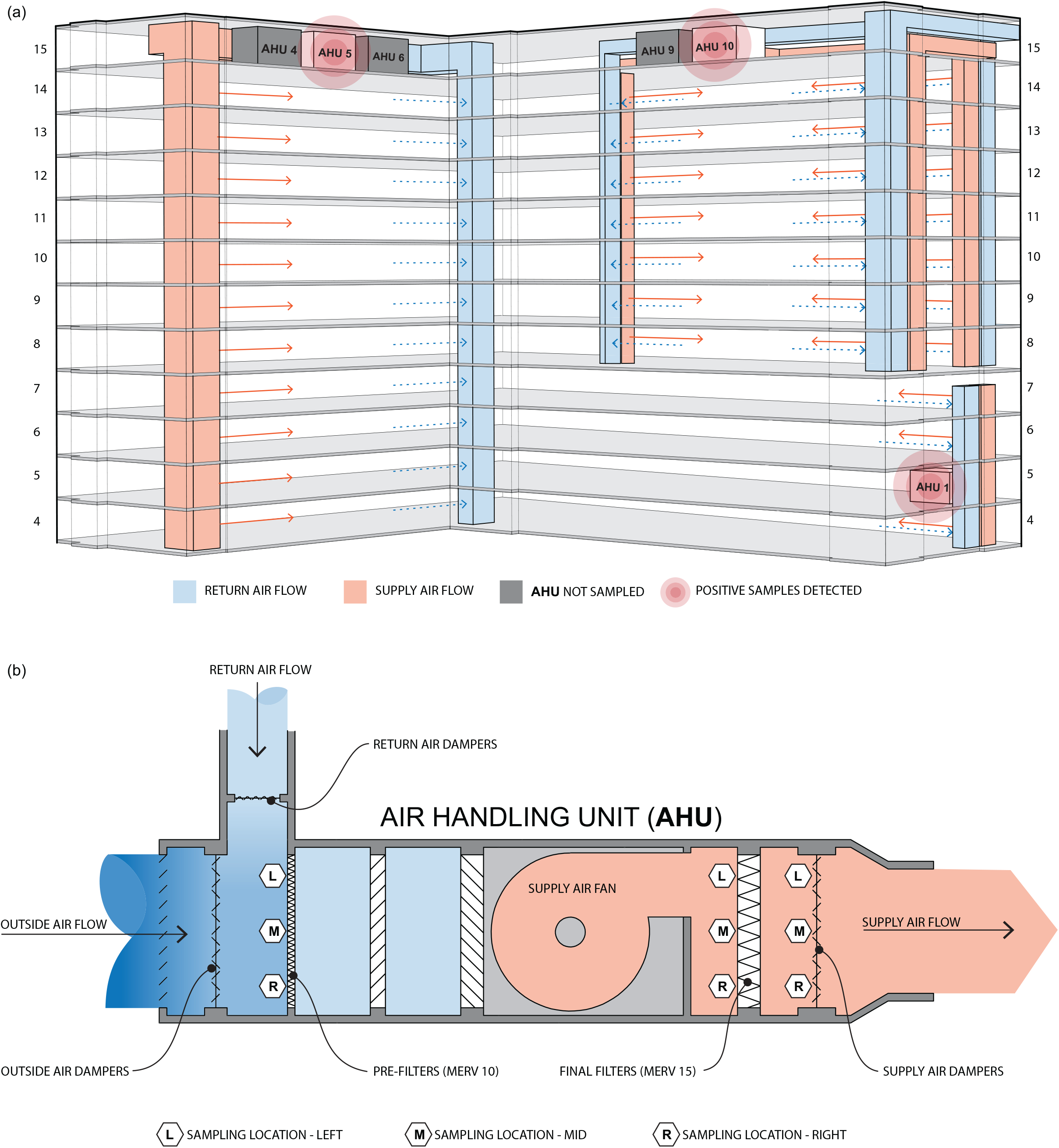
(a) System diagram of OHSU hospital ventilation supply and return air collected and distributed to multiple floors by sampled AHUs. (b) AHU sectional diagram illustrating the path of airflow, mixing of recirculated return air with outdoor air fraction and locations of swab sampling.

### Sample Processing and Molecular Analysis

Samples were hand-carried to a research laboratory at OHSU for initial processing. In a class 2 biosafety cabinet (BSC), conical tubes were vortexed briefly, allowed to settle for five minutes, and 200 µL of the supernatant was removed and combined with 600 µL of the lysis/preservative buffer (DNA/RNA Shield, Zymo Research). Samples were then transported by car to a BSL-2 laboratory at the University of Oregon campus in Eugene, Oregon, USA. Total RNA was extracted from all samples using Zymo Quick-DNA/RNA Viral MagBead kit (Zymo Research #R2141) and stored at −80°C until analysis. The abundance of SARS-CoV-2 in each sample was determined using quantitative reverse-transcription polymerase chain reaction (qRT-PCR) targeting a 157 bp segment of the SARS-CoV-2 spike glycoprotein gene^4^. An artificial gene standard from Integrated DNA Technologies with known copy number was utilized to create a dilution series and standard curve for the determination of viral gene copies in each sample^23^, with a limit of detection of 11.1 gene copies. All qRT-PCR reactions were run in triplicate and any gene copies observed in no template controls were removed from samples in each plate to account for interrun variability. Potential contamination during initial processing was assessed through the use of passive air settling plates and reagent controls within the BSL2 lab. To accomplish this, passive air settling plates were placed in the BSC and on the outside lab bench for the duration of processing. Following the completion of specimen processing, the same swabs, viral transport media, and processing steps were performed in an identical fashion to environmental samples for the controls. Reagent controls were processed concurrently with environmental samples. All controls tested negative for the presence of SARS-CoV-2 RNA.

## Results

In total, 56 samples from three different AHUs were collected; 25% (14/56) of samples contained detectable SARS-CoV-2 RNA (Table S1). The highest abundance sample (∼245 gene copies) was found on the pre-filters, where outside air mixes with recirculated building air. Of the samples collected, 35% (7/20) of samples at the pre-filters, 16.67% (2/12) of samples at the final filter, and 20.8% (5/24) of samples at the supply air dampers contained detectable SARS-CoV-2 RNA (Table 1). The least SARS-CoV-2 RNA was detected at the final filter and the most at the pre-filters (Figure 2).

**Table 1.** Summary statistics and percent of positive samples from each sampling location within all AHUs.

|  | Pre-Filters |  | Final Filters |  | Supply Air Dampers |  |
| --- | --- | --- | --- | --- | --- | --- |
|  | Total Number (n) | Number Positive (%) | Total Number (n) | Number Positive (%) | Total Number (n) | Number Positive (%) |
|  | 20 | 7 (35) | 12 | 2 (16.67) | 24 | 5 (20.8) |
| Cumulative Gene Copies (X) | 354.8 (34.2) |  | 103.2 (86.2) |  | 342.5 (77.7) |  |

**Figure 2:**
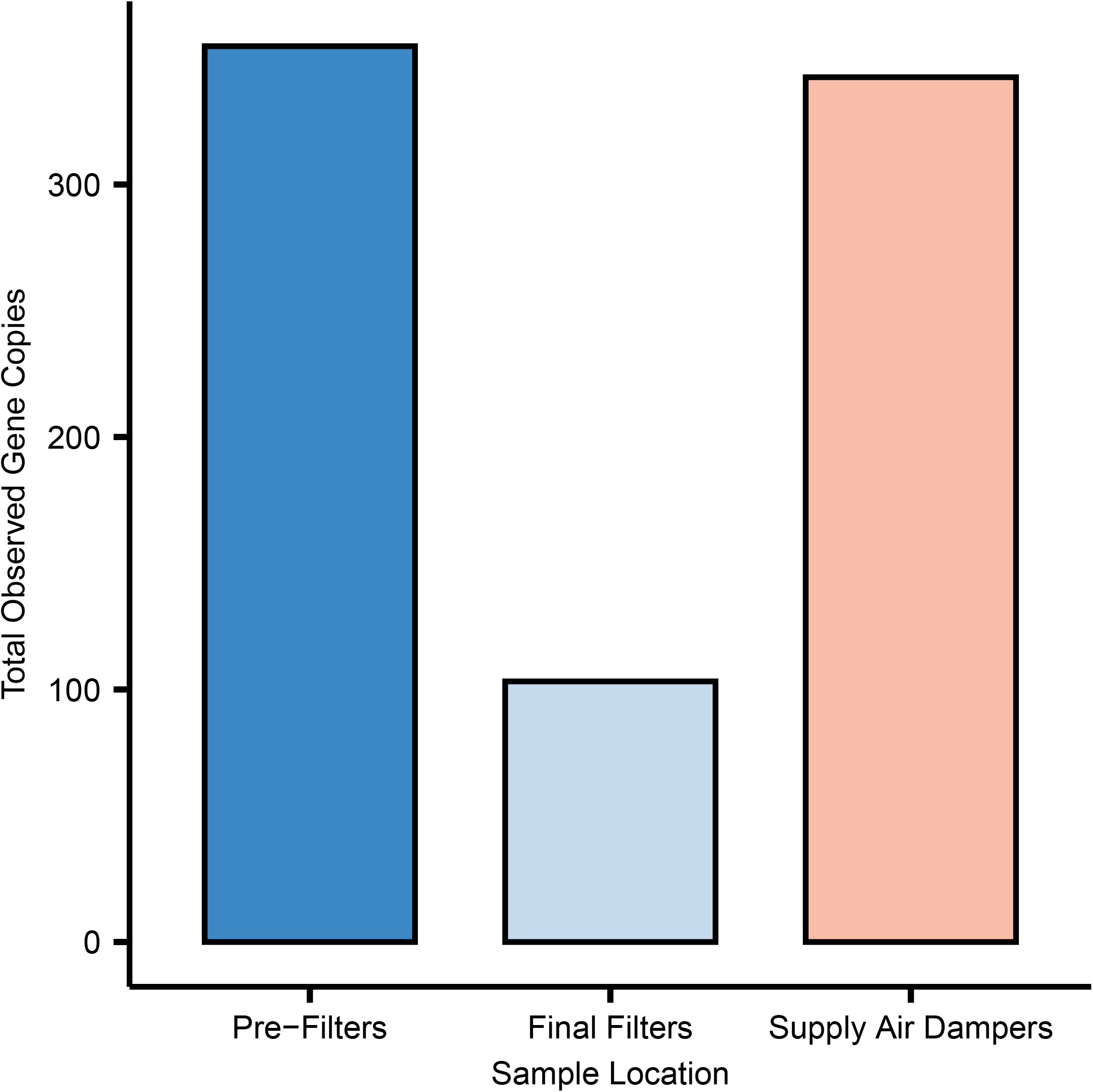
Observed total number of SARS-CoV-2 gene copies at the pre-filters (left), final filters (middle), and supply air dampers (right).

## Discussion

This investigation demonstrates the presence of SARS-CoV-2 RNA at multiple locations within mechanical AHUs, and more specifically, AHUs serving multiple floors of a hospital tower in which COVID-19 patients were housed. Furthermore, SARS-CoV-2 RNA remained detectable 33.3% (5/15) of the time at the final sampling location (supply air damper), after the recirculated air had been mixed with fresh outside air, passed through the pre-filter (MERV 10) and final filter (MERV15) stages. This suggests that the filtration practices in place in some of the most highly filtered environments, such as healthcare, does not eliminate the passage of SARS-CoV-2 viral RNA, and potentially SARS-CoV-2 viral particles, through HVAC systems and potentially back into the supply air. The infectious potential of this viral genetic material is currently unknown. These data demonstrate the potential that air evacuated from building spaces containing infectious individuals may be recirculated and distributed to other building spaces through centralized HVAC systems while containing SARS-CoV-2 RNA (and possibly virus), even after the filtration process and the dilution from the addition of 70-80% outside air, (thus, only 20-30% recirculated air, during the sampling period).

Although positive samples were not assessed for infectivity of SARS-CoV-2, the presented data supports previous claims of aerosolized viral particles^8,10–12^ that may be carried remotely from their source by indoor air currents produced by built environment HVAC systems. Despite the growing evidence of the potential airborne nature of SARS-CoV-2 and the potential for aerosolized particles to contribute to some transmission events,^6–8,10–12,24,25^, little guidance is given by both the World Health Organization^26^ (WHO) and Centers for Disease Control^27^ (CDC) concerning the potential for viral spread through airborne routes of transmission. The American Society of Heating Refrigerating and Air-Conditioning Engineers (ASHRAE) Standard 170-2017 Ventilation for Healthcare Facilities guidelines requires MERV 7 minimum for filter bank one and MERV 14 minimum for filter bank two in healthcare settings such as those studied here^22^. The building studied herein exceeds requirements for filtration efficiency at both filter bank locations. ASHRAE’s current Position Document on Infectious Aerosols encourages consideration of HEPA filtration in healthcare buildings and MERV 13 or higher in non-healthcare buildings^28^.

Placement of patients with known or suspected COVID-19 in negative pressure rooms when available, as recommended by CDC, greatly reduces the risk of re-circulation of virus particles within HVAC systems. However, most hospitals and outpatient clinics do not have sufficient numbers of negative pressure rooms to accommodate all patients with known or suspected COVID-19. Furthermore, a substantial number of infected individuals that are asymptomatic or pre-symptomatic can shed aerosolized viral particles to shared air spaces. This risk of transmission may be even higher in non-healthcare buildings that often do not have qRT-PCR based screening practices, and typically have lower air exchange rates and less efficient filtration, and during extreme weather conditions when HVAC thermal system capacities cannot manage thermal comfort with higher outside air fractions.

Previous studies have demonstrated that SARS-CoV-2 can be found in aerosols and droplets ranging from 0.25-4 microns^10,11^. In experimentally generated aerosols, SARS-CoV-2 has been demonstrated to retain infectivity for between one and sixteen hours^29–32^, lending credence to the potential for aerosolized transmission to occur.

There are steps that can be taken to limit the potential impact of airborne dissemination of viruses in the built environmental, including careful donning and doffing of personal protective equipment^15^, hand hygiene after hand contact with the environment, cleaning of high touch surfaces, and use of UV radiation^33^. Use of the highest possible efficiency filters for the building type and HVAC system design^6^, ensuring indoor relative humidity is between 40%-60%^30^, and increasing the fraction of outside air introduced into indoor environments^6^ are additional strategies that could mitigate transmission of virus through ventilation systems.

There were several limitations to this study. First, samples were not evaluated for the presence of viable SARS-CoV-2 virus. Second, to prevent disruption of hospital operations, routine sampling of all AHUs was not possible, limiting temporal analysis. Third, the sample point of a filter or damper can only be representative of that sampling area and the porous nature of the filters may inhibit efficient specimen recovery. Lastly, this is a focused examination of one specific exposure factor and does not address several others (exposure routes, sampling conditions, viability) and should therefore only serve as part of the equation in understanding the overall exposure risk of SARS-CoV-2.

## Conclusion

This study demonstrates SARS-CoV-2 RNA contamination throughout several AHUs path of flow, including return air, two filtration stages, and supply air, for multiple floors of the hospital and serves as the first evidence of the potential for SARS-CoV-2 RNA (and possibly virus), irrespective of viability, to enter into and travel throughout HVAC systems. While there is still a paucity of information on the potential viability and infectivity of the present SARS-CoV-2, this paper demonstrates that actions to protect against the potential for SARS-CoV-2 aerosolized travel, and subsequent transmission, should be taken into account in built environment mitigation strategies. Specifically, the data suggest that actions should be taken by healthcare facilities immediately in order to avoid or minimize potential future SARS-CoV-2 healthcare associated infection given that risks increase as more extreme weather conditions force a reduction in the outside air admitted to buildings.

## Data Availability

All data available upon request.

## Acknowledgements

The authors would like to thank Oregon Health and Science University for help in collecting samples from AHUs. The authors would like to thank Georgia MacCrone for her help in processing samples. The authors would like to thank the University of Oregon Genomics and Cell Characterization Core Facility (GC3F) for their expertise and resources. The authors would like to thank Flic Coulter and Corinne Fargo from the Messer Laboratory for their contributions in sample processing and sampling collection method input.

## References

1. Coronavirus. https://www.who.int/emergencies/diseases/novel-coronavirus-2019. Accessed June 21, 2020.

2. Galbadage T, Peterson BM, Gunasekera RS. Does COVID-19 Spread Through Droplets Alone? Front Public Health. 2020;8:163. April 24, 2020. http://dx.doi.org/10.3389/fpubh.2020.00163.

3. Miller SL, Nazaroff WW, Jimenez JL, et al. Transmission of SARS-CoV-2 by inhalation of respiratory aerosol in the Skagit Valley Chorale superspreading event. Infectious Diseases (except HIV/AIDS). June 2020. June 17, 2020. https://www.medrxiv.org/content/10.1101/2020.06.15.20132027v2.abstract.

4. Chan JF-W, Yuan S, Kok K-H, et al. A familial cluster of pneumonia associated with the 2019 novel coronavirus indicating person-to-person transmission: a study of a family cluster. Lancet. 2020;395:514–523. February 15, 2020. http://dx.doi.org/10.1016/S0140-6736(20)30154-9.

5. Modes of transmission of virus causing COVID-19: implications for IPC precaution recommendations. https://www.who.int/news-room/commentaries/detail/modes-of-transmission-of-virus-causing-covid-19-implications-for-ipc-precaution-recommendations. Accessed June 22, 2020.

6. Allen JG, Marr LC. Recognizing and controlling airborne transmission of SARS □ CoV □ 2 in indoor environments. Indoor Air. 2020;30:557–558. 2020. http://dx.doi.org/10.1111/ina.12697.

7. Shin Young Park, Young-Man Kim, Seonju Yi, et al. Coronavirus Disease Outbreak in Call Center, South Korea. Emerging Infectious Disease journal. 2020;26. 2020. https://www.nc.cdc.gov/eid/article/26/8/20-1274_article.

8. Jianyun Lu, Jieni Gu, Kuibiao Li, et al. COVID-19 Outbreak Associated with Air Conditioning in Restaurant, Guangzhou, China, 2020. Emerging Infectious Disease journal. 2020;26. 2020. https://www.nc.cdc.gov/eid/article/26/7/20-0764_article.

9. Ong SWX, Tan YK, Chia PY, et al. Air, Surface Environmental, and Personal Protective Equipment Contamination by Severe Acute Respiratory Syndrome Coronavirus 2 (SARS-CoV-2) From a Symptomatic Patient. JAMA. 2020. 2020. http://dx.doi.org/10.1001/jama.2020.3227.

10. Santarpia JL, Rivera DN, Herrera V, et al. Transmission Potential of SARS-CoV-2 in Viral Shedding Observed at the University of Nebraska Medical Center. Infectious Diseases (except HIV/AIDS). March 2020. March 26, 2020. https://www.medrxiv.org/content/10.1101/2020.03.23.20039446v2.

11. Liu Y, Ning Z, Chen Y, et al. Aerodynamic analysis of SARS-CoV-2 in two Wuhan hospitals. Nature. April 2020. April 27, 2020. http://dx.doi.org/10.1038/s41586-020-2271-3.

12. Correia G, Rodrigues L, Gameiro da Silva M, Gonçalves T. Airborne route and bad use of ventilation systems as non-negligible factors in SARS-CoV-2 transmission. Med Hypotheses. 2020;141:109781. August 2020. http://dx.doi.org/10.1016/j.mehy.2020.109781.

13. Singh R. Split Air Conditioners and their role in Airborne Infection Spread: Short Communication/Research Note. IndiaRxiv. April 2020. April 7, 2020. https://indiarxiv.org/jp9te/download?format=pdf.

14. Dai H, Zhao B. Association of infected probability of COVID-19 with ventilation rates in confined spaces: a Wells-Riley equation based investigation. Emergency Medicine. April 2020. April 24, 2020. http://dx.doi.org/10.1101/2020.04.21.20072397.

15. Zhang R, Li Y, Zhang AL, Wang Y, Molina MJ. Identifying airborne transmission as the dominant route for the spread of COVID-19. Proc Natl Acad Sci U S A. June 2020. June 11, 2020. http://dx.doi.org/10.1073/pnas.2009637117.

16. Fantini MP, Reno C, Biserni GB, Savoia E, Lanari M. COVID-19 and the re-opening of schools: a policy maker’s dilemma. Italian Journal of Pediatrics. 2020;46. 2020. http://dx.doi.org/10.1186/s13052-020-00844-1.

17. Qian H, Zheng X. Ventilation control for airborne transmission of human exhaled bioaerosols in buildings. J Thorac Dis. 2018;10:S2295–S2304. July 2018. http://dx.doi.org/10.21037/jtd.2018.01.24.

18. Menzies D, Fanning A, Yuan L, FitzGerald JM. Hospital ventilation and risk for tuberculous infection in canadian health care workers. Canadian Collaborative Group in Nosocomial Transmission of TB. Ann Intern Med. 2000;133:779–789. November 21, 2000. http://dx.doi.org/10.7326/0003-4819-133-10-200011210-00010.

19. Lutz BD, Jin J, Rinaldi MG, Wickes BL, Huycke MM. Outbreak of invasive Aspergillus infection in surgical patients, associated with a contaminated air-handling system. Clin Infect Dis. 2003;37:786–793. September 15, 2003. http://dx.doi.org/10.1086/377537.

20. Atkinson J, Chartier Y, Pessoa-Silva CL, Jensen P, Li Y, Seto W-H. Infection and Ventilation. World Health Organization; 2009. 2009. https://www.ncbi.nlm.nih.gov/books/NBK143278/. Accessed June 22, 2020.

21. Schoen BYLJ, E. P, FELLOW/LIFE MEMBER ASHRAE. Guidance for Building Operations During the COVID-19 Pandemic. https://www.uc.edu/content/dam/uc/af/facilities/fme/20%20ASHRAE%20SARS-CoV-2%20Guidance.pdf.

22. ANSI/ASHRAE/ASHE Standard 170-2017, Ventilation of Health Care Facilities. https://www.ashrae.org/technical-resources/standards-and-guidelines/standards-addenda/ansi-ashrae-ashe-standard-170-2017-ventilation-of-health-care-facilities. Accessed June 25, 2020.

23. Fahimipour AK, Hartmann EM, Siemens A, et al. Daylight exposure modulates bacterial communities associated with household dust. Microbiome. 2018;6:175. October 18, 2018. http://dx.doi.org/10.1186/s40168-018-0559-4.

24. DirkJan Hijnen, Angelo Valerio Marzano, Kilian Eyerich, et al. SARS-CoV-2 Transmission from Presymptomatic Meeting Attendee, Germany. Emerging Infectious Disease journal. 2020;26. 2020. https://www.nc.cdc.gov/eid/article/26/8/20-1235_article.

25. Hamner L. High SARS-CoV-2 attack rate following exposure at a choir practice—Skagit County, Washington, March 2020. MMWR Morb Mortal Wkly Rep. 2020;69. 2020. https://www.cdc.gov/mmwr/volumes/69/wr/mm6919e6.htm?utm_medium=email&utm_source=govdelivery.

26. Organization WH, Others. Infection Prevention and Control during Health Care When COVID-19 Is Suspected: Interim Guidance, 19 March 2020. World Health Organization; 2020. 2020. https://apps.who.int/iris/bitstream/handle/10665/331495/WHO-2019-nCoV-IPC-2020.3-eng.pdf.

27. CDC. Coronavirus Disease 2019 (COVID-19) – Prevention & Treatment. Centers for Disease Control and Prevention. June 12, 2020. https://www.cdc.gov/coronavirus/2019-ncov/prevent-getting-sick/prevention.html. Accessed June 22, 2020.

28. Stewart EJ, Schoen LJ. The ASHRAE Position Document on Infectious Aerosols was developed by the Society’s Environmental Health. https://www.ashrae.org/file%20library/about/position%20documents/pd_infectiousaerosols_2020.pdf.

29. Alyssa C. Fears, William B. Klimstra, Paul Duprex, et al. Persistence of Severe Acute Respiratory Syndrome Coronavirus 2 in Aerosol Suspensions. Emerging Infectious Disease journal. 2020;26. 2020. https://www.nc.cdc.gov/eid/article/26/9/20-1806_article.

30. Casanova LM, Jeon S, Rutala WA, Weber DJ, Sobsey MD. Effects of air temperature and relative humidity on coronavirus survival on surfaces. Appl Environ Microbiol. 2010;76:2712–2717. May 2010. http://dx.doi.org/10.1128/AEM.02291-09.

31. Sizun J, Yu MW, Talbot PJ. Survival of human coronaviruses 229E and OC43 in suspension and after drying onsurfaces: a possible source ofhospital-acquired infections. J Hosp Infect. 2000;46:55–60. September 2000. http://dx.doi.org/10.1053/jhin.2000.0795.

32. van Doremalen N, Bushmaker T, Morris DH, et al. Aerosol and Surface Stability of SARS-CoV-2 as Compared with SARS-CoV-1. N Engl J Med. March 2020. March 17, 2020. http://dx.doi.org/10.1056/NEJMc2004973.

33. Keil SD, Ragan I, Yonemura S, Hartson L, Dart NK, Bowen R. Inactivation of severe acute respiratory syndrome coronavirus 2 in plasma and platelet products using a riboflavin and ultraviolet light-based photochemical treatment. Vox Sang. April 2020. April 20, 2020. http://dx.doi.org/10.1111/vox.12937.

